# Malaria Incidence Rose Following the introduction of Neonicotinoid-Based IRS in selected districts in northern Ghana: An Observational Analysis

**DOI:** 10.1101/2025.09.16.25335957

**Authors:** Sylvester Coleman, Christian Atta-Obeng, Clinton Nkolokosa, Abdul Gafaru Mohammed, Otubea Owusu Akrofi, Ihsan Isaka, Wahjib Mohammed, Nana Yaw Peprah, Samuel Asiedu, Samuel K. Dadzie, Dominic B. Dery, Julie-Anne A. Tangena, Adrienne Epstein, Keziah Malm

## Abstract

In Ghana, indoor residual spraying (IRS) has been a key intervention for malaria control since 2008. After seven years of IRS with an organophosphate insecticide, which substantially reduced malaria incidence, IRS programs transitioned to neonicotinoid-based products between 2018 and 2019 as part of an insecticide rotation plan. This change was informed largely by entomological data from limited pilot districts, with little evidence on epidemiological impact. We assessed the effect of this transition using monthly, district-level malaria incidence from 2015-2022 in 22 IRS districts across four northern regions. Incidence was calculated from routinely reported confirmed malaria cases in the Ghana Health Service District Health Information Management System, adjusted for testing rates, with population denominators from census-based estimates. An interrupted time series model with generalized estimating equations was fit to 2015-2019 data, when organophosphates were in use, to generate counterfactual trends through 2022, adjusting for rainfall, temperature, vegetation index, seasonality, and initiation of seasonal malaria chemoprevention. Observed incidence exceeded counterfactual estimates in the first transmission season after the switch and remained higher in subsequent campaigns. Across all districts, incidence was 26% higher under neonicotinoid IRS than expected under continued organophosphate use (IRR = 1.26, 95% CI 1.07-2.08). Regional differences were observed, with significant increases in the Northern (IRR = 1.76, 95% CI 1.04-2.74), North East (IRR = 1.83, 95% CI 1.08-3.40), and Upper East (IRR = 4.26, 95% CI 2.17-8.50) regions, but not in the Upper West (IRR = 1.25, 95% CI 0.90-1.70). These findings suggest that the transition to neonicotinoids may have reduced IRS effectiveness in northern Ghana. Future pilots of new IRS products should incorporate both entomological and epidemiological outcomes to guide region-specific selection before national scale-up.

## Introduction

Indoor residual spraying (IRS) is an important tool for the control and elimination of malaria [1, 2]. Increased investments in malaria control in the last two decades, including IRS coverage, has permitted an expansion of vector control measures leading to an estimated 37% decline in malaria incidence globally [3, 4]. These gains have been reported across diverse geographical landscapes and against various mosquito vectors [5–9]. However, since 2018, the rate of decline in malaria burden has slowed in most countries, and some have even reported an increase in cases [10, 11]. In Ghana, progress in reducing malaria cases has either stalled or reversed, with an estimated 5.3 million cases in 2022 compared to 5.2 million in 2018 [11].

A major challenge in reducing malaria is maintaining the efficacy of vector control tools, which is increasingly threatened by the complex and ever evolving resistance of mosquitoes to public health insecticides [12]. A direct consequence of resistance development in malaria vectors to existing control tools has been the need to shift towards insecticide products with alternative chemistries and modes of action [13]. Historically, vector control programs relied excessively on pyrethroids and carbamates for IRS [14]. However, IRS programs shifted to organophosphates in 2012 with the introduction of the microencapsulated formulation of Actellic 300CS (active ingredient [a.i.] pirimiphos-methyl) [15, 16], and subsequently to neonicotinoids with the introduction of SumiShield 50WG (a.i. clothianidin) in 2018 and Fludora Fusion (a.i. clothianidin and deltamethrin) in 2020 [17, 18]. In 2022, 11 out of the 13 sub-Saharan African countries supported by the President’s Malaria Initiative (PMI) to implement IRS in their vector control programmes included neonicotinoids in their strategies [19]. Yet, there is still limited evidence supporting the effectiveness of these WHO prequalified neonicotinoid-based formulations in suppressing malaria incidence, with initial field studies showing mixed results [20–22].

In Ghana, IRS has been a key component of the malaria control strategy since 2008 [23, 24]. Whereas insecticide treated nets (ITNs) have been distributed widely throughout Ghana, IRS has been centred in selected districts in northern Ghana where malaria burden is highest [25] and in Obuasi municipal and Obuasi East Districts in the Ashanti region, where the AngloGold Ashanti Gold mine supports vector control activities. Since the switch of IRS from pyrethroids to organophosphate formulations in 2012, with carbamates (Propoxur) used only in 2013, organophosphates (pirimiphos-methyl) remained the sole IRS insecticide until 2018, when neonicotinoids were introduced and gradually adopted [26]. A review on the introduction of non-pyrethroid based insecticides (primarily pirimiphos-methyl CS) in IRS programs across Africa revealed that the transition helped maintain program efficacy [27, 28]. In Ghana, an observational analysis of routine health data demonstrated a significant reduction in malaria incidence in districts that switched IRS from pyrethroids to organophosphates [7]. In 2018, neonicotinoid-based formulations were introduced in Ghana and had fully replaced organophosphates by 2021. This transition aligned with Ghana’s insecticide resistance management plan, which promotes the rotation of insecticides with different modes of action to slow the development of resistance. As part of this strategy, the IRS program under the National Malaria Elimination Program (NMEP) piloted SumiShield® 50WG in five districts. Based on residual efficacy results from this pilot and susceptibility data from multiple districts, neonicotinoid-based insecticides were adopted for broader IRS implementation starting in 2019. Although understanding the impact of this transition on malaria incidence is important, no epidemiological impact study has been conducted in Ghana to date. In this study, we assessed the impact of neonicotinoid-based insecticides on malaria incidence using monthly health facility data from 2015 to 2022 across 22 districts in northern Ghana.

## Materials and methods

### Study area

The study area comprises 22 districts in northern Ghana where IRS implementation shifted from organophosphate to neonicotinoid-based insecticides between 2018 and 2020 (Fig 1). These districts are located in the northern Savannah zone (comprised of the Guinea and Sudan Savannah), which is characterized by a relatively dry climate, with a unimodal rainy season that begins in May and ends in October, and precipitation ranges between 750 mm and 1050 mm annually [29]. Across the study area, elevation ranges from approximately 100 to 500 m above sea level, with land cover mainly dominated by savanna woodland interspersed with cultivated cropland, settlement, and water bodies [30, 31].

**Fig 1.**
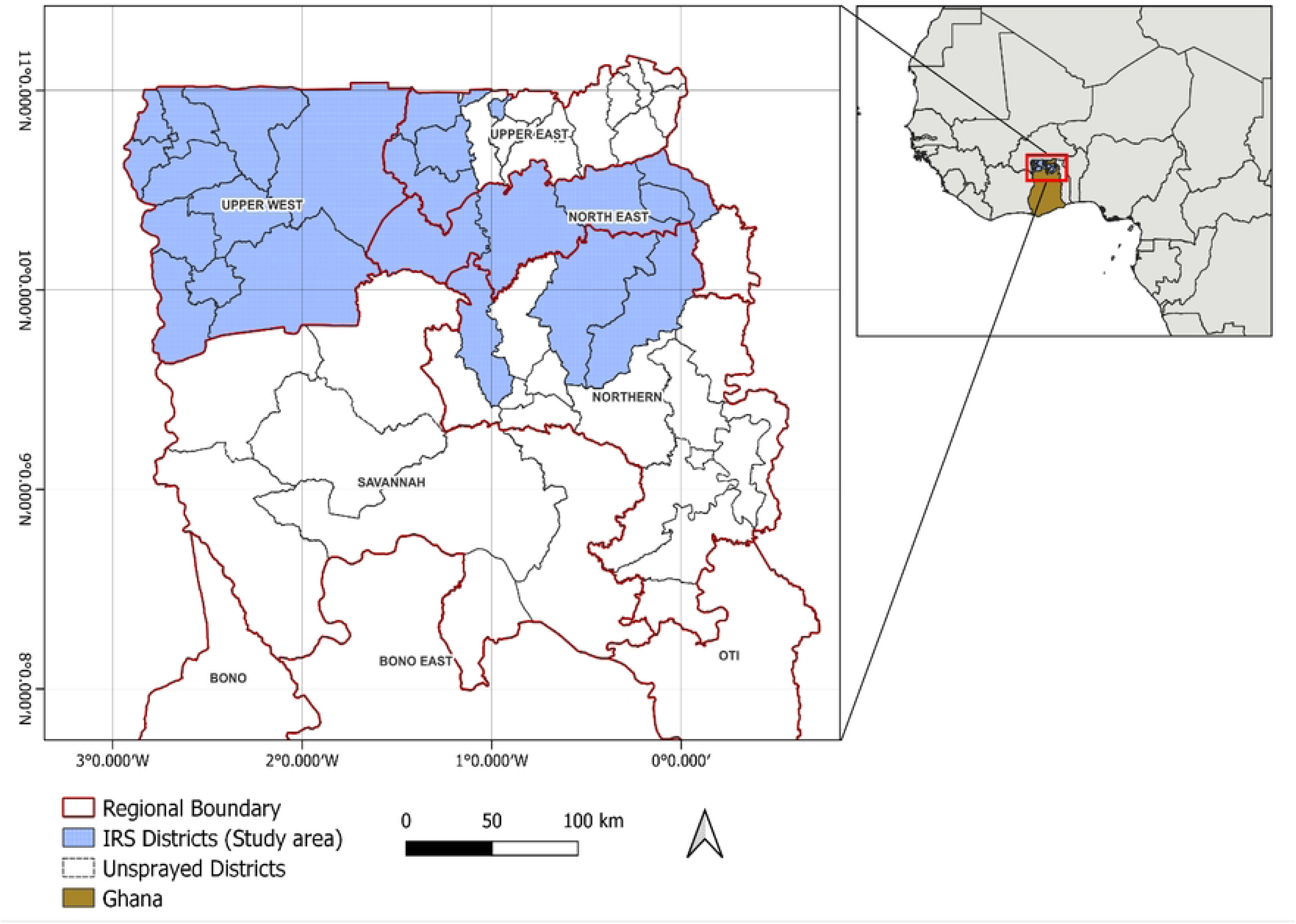
Map of Ghana showing the study area (IRS districts -blue shading) across Upper West, Upper East, North East, and Northern region of Ghana. *Red boundaries indicate regional administrative borders. The inset highlights Ghana’s location within West Africa*.

Malaria is endemic and seasonal in the northern Savannah zone with peak malaria transmission occurring between July and November [32]. The main vectors in the study area are *An. gambiae*, *An. coluzzii* and *An. arabiensis*. Resistance assays in the study area indicate high levels of pyrethroid resistance, while vectors have remained largely susceptible to both pirimiphos-methyl and clothianidin since their introduction [19]. An exception was observed in East Mamprusi District, where resistance to pirimiphos-methyl was detected in 2019, but susceptibility returned in subsequent years [19, 33].

Vector control in our study area consists of IRS with ITNs primarily distributed to pregnant women and children below five years through antenatal or child welfare clinics [34]. Additionally, intermittent preventive treatment of malaria in pregnancy IPTp-SP policy was adopted in 2003 as a national policy [35]. Seasonal malaria chemoprevention (SMC) was later introduced, beginning in the Upper West Region in 2015, followed by scale-up to the Upper East Region in 2016, and subsequently to the Northern and North East Regions in 2019. Since 2019, SMC has been implemented across all IRS districts as part of the broader malaria control strategy[36].

IRS in the northern region was introduced in 2008 in four districts and until recently, with funding from PMI and the Global Fund, a total of 23 districts were sprayed annually. This included 11 districts in Upper West, five of six districts in North East, three of 15 in Upper East, and four of 17 in Northern Region. Thus, our study districts comprise 22 of the 23 IRS districts (Fig 1), with the exception of Tatale-Sangule, which has never received organophosphate-based IRS. These districts have historically been targeted for IRS because they were classified as areas of high malaria transmission.

A summary of insecticides used across study districts is provided in Supplemental S1 Table, with formulation details in Table 1. Most districts in the Northern and North East regions began IRS between 2008 and 2013, initially with pyrethroids, before switching to the organophosphate Actellic® 300CS (AC) from 2012. In Upper East and Upper West, IRS started in 2012 with VectoGuard (a pirimiphos-methyl formulation), shifted to the carbamate ProGuard (propoxur) in 2013, and to Actellic® 300CS in 2014. From 2018 onwards, there was a progressive shift to neonicotinoids: SumiShield® 50WG (SS) and later Fludora® Fusion (FF). An exception was in Mamprugu-Moagduri district, where spraying moved from Actellic to SumiShield, back to Actellic, and then to neonicotinoids (SS and FF). Monthly wall cone bioassays consistently showed that all IRS formulations maintained residual efficacy for over ten months. Actellic® 300CS (pirimiphos-methyl) remained effective for up to 11 months, while Fludora® Fusion and SumiShield®50WG (clothianidin-based) also remained effective for at least 11 months across all surface types [37]. These residual profiles are sufficient to provide protection throughout the local malaria transmission season, typically lasting six-to-seven-months [38].

**Table 1.** Type of IRS insecticide formulation used in our study districts from 2015-2022.

| Region | Districts | IRS start year | 2015 | 2016 | 2017 | 2018 | 2019 | 2020 | 2021 | 2022 |
| --- | --- | --- | --- | --- | --- | --- | --- | --- | --- | --- |
| North East | Bunkpurugu - Nakpanduri | 2011 | AC | AC | AC | AC | AC | SS | SS | FF |
|  | East Mamprusi | 2009 | AC | AC | AC | AC | SS | FF | SS | SS |
|  | Mamprugu-Moagduri | 2008 | AC | AC | AC | SS | SS | AC | FF | SS |
|  | West Mamprusi | 2008 | AC | AC | AC | AC | SS | SS | FF | FF |
|  | Yunyoo-Nasuan | 2011 | AC | AC | AC | AC | AC | FF | SS | SS |
| Northern | Gushiegu | 2008 | Nsp | Nsp | AC | AC | AC | FF | SS | SS |
|  | Karaga | 2008 | Nsp | Nsp | AC | AC | AC | SS | SS | FF |
|  | Kumbungu | 2008 | AC | AC | AC | AC | AC | FF | FF | SS |
| Upper East | Builsa North | 2013 | Nsp | Nsp | AC | AC | SS | SS | SS | FF |
|  | Builsa South | 2013 | Nsp | Nsp | AC | AC | SS | SS | SS | FF |
|  | Kasena-Nankana West | 2013 | Nsp | Nsp | AC | AC | SS | SS | SS | FF |
| Upper West | Daffiama-Bussie-Issa | 2012 | AC | AC | AC | SS | SS | FF | FF | SS |
|  | Jirapa | 2012 | AC | AC | AC | SS | SS | FF | FF | SS |
|  | Lambussie | 2012 | AC | AC | AC | AC | SS | SS | SS | FF |
|  | Lawra | 2012 | AC | AC | AC | AC | SS | SS | SS | FF |
|  | Nadowli-Kaleo | 2012 | AC | AC | AC | SS | SS | FF | SS | SS |
|  | Nandom | 2012 | AC | AC | AC | AC | SS | SS | SS | FF |
|  | Sissala East | 2013 | AC | AC | AC | AC | SS | SS | SS | FF |
|  | Sissala West | 2012 | AC | AC | AC | SS | SS | FF | SS | FF |
|  | Wa East | 2012 | AC | AC | AC | AC | SS | SS | SS | FF |
|  | Wa Municipal | 2012 | AC | AC | AC | AC | SS | FF | FF | SS |
|  | Wa- West | 2012 | AC | AC | AC | AC | SS | SS | FF | FF |

Blue AC- the organophosphate Actellic® 300CS (pirimiphos methyl), gold SS- the neonicotinoid SumiShield™ 50WG (Clothianidin only), green FF- the neonicotinoid Fludora® Fusion WP SB (Clothianidin + Deltamethrin). NSp=not sprayed.

### Health-facility and environmental data

The primary outcome for this analysis is district-level malaria incidence recorded per 1000 person-years, measured through data collected routinely from health facility malaria cases, adjusted for the testing rate. We estimated the malaria incidence using validated monthly passive malaria case data reported from 2015 to 2022 in the District Health Information Management System (DHIS2) of the Ghana Health Service. These data, compiled by district, encompass the number of suspected malaria cases, tested suspected malaria cases, and confirmed malaria cases identified via microscopy or rapid diagnostic tests. Population values were estimated from the 2021 Population and Housing Census in DHIS2 (District Health Information Software 2), adjusted for estimated annual growth. Data from the 2022 Demographic and Health Survey indicate that the proportion of febrile children for whom care was sought is notably high across the study area, with reported coverage of 75.3% in Upper West, 74.8% in Upper East, 71.7% in North East, and 62.2% in the Northern Region [39].

The period 2015–2022 was selected for this analysis because of the improved data quality assurance since 2015. DHIS2 data have historically been subject to quality concerns, including inconsistent reporting and delayed data entry. Analyses of malaria surveillance data from 2014– 2017 have confirmed improved reliability of DHIS2 entries, particularly in IRS implementation areas [7]. Validation exercises conducted at facility level and supervised by District Health Management Teams have resulted in documented improvements in data completeness and internal consistency, with accuracy of c. 90% [40].

We adjusted for time-varying environmental variables that are associated with changes in malaria incidence, averaged at the district level. These include minimum and maximum monthly temperature in Celsius degrees [41], precipitation [42], NDVI [31], an indicator variable for calendar month (to adjust for seasonal effects), and an indicator variable representing whether the district had initiated SMC.

This work was conducted with permission and approval from the National Malaria Elimination Program as part of their mandate to review malaria intervention impact in Ghana.

### Statistical analysis

An interrupted time series (ITS) approach [43, 44] was taken using R software and the *geepack* package [45] to assess the impact of the change in organophosphate to neonicotinoid-based IRS on malaria incidence. The following segmented regression model was specified:

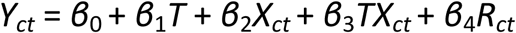

where *Y_ct_* is the outcome (malaria incidence) in district *c* at month *t*, *T* is the time elapsed since the start of the study in months, *X_ct_* is a dummy variable indicating the organophosphate-based IRS period (0) or neonicotinoid-based IRS period (1) for district *c* at month *t*, and *R_ct_* is the vector of covariates for district *c* at month *t.* As such, *β*_0_ represents malaria incidence at the start of the study (t=0), *β*_1_ represents the change in malaria incidence associated with a one-month increase in the organophosphate-based IRS period, *β*_2_ represents the level change in malaria incidence in the neonicotinoid-based IRS period, and *β*_3_ represents the additional change in the slope after the change in IRS active ingredients. Generalized estimating equations were used to account for clustering at the district level. We used a Poisson distribution and modelled the count of malaria cases in district *c* at month *t*, with an offset of the logged population denominator and an autoregressive order of 1 correlation structure to account for serial autocorrelation. The resulting segmented ITS model was used to estimate the counterfactual (unobserved) trend of malaria incidence in the absence of a change in IRS active ingredients (assuming that organophosphate-based IRS was continued) for each month by setting *X_ct_* to zero. Incidence rate ratios (IRR) were calculated by comparing the observed incidence to the counterfactual incidence, with bootstrapped 95% confidence intervals (CI). Models were first estimated across all districts. Subsequently, secondary analyses allowed the post-intervention slope change to vary by region by including a three-way interaction term between intervention, time since intervention, and region. This was motivated by the fact that in Ghana, health system structure and the delivery of IRS and other malaria control interventions are organized at the regional level, which could plausibly lead to regional differences in the temporal pattern of impact. We conducted an additional analysis to determine whether the impact of the change in the IRS active ingredient differed across transmission intensity (low transmission < 200 cases per 1,000 person-years during the baseline period; moderate transmission >= 200 cases per 1,000 person-years during the baseline period).

Finally, we conducted a sensitivity analysis to determine whether results differed between the two clothianidin-based IRS products (SumiShield vs. Fludora®Fusion). We re-fit the interrupted time series model replacing the single neonicotinoid-IRS based indicator with a categorical variable for insecticide type (Actellic, SumiShield, Fludora®Fusion) and included a time-since-spray variable that reset at the start of each IRS round. This allowed estimation of separate immediate level changes and post-spray slopes for each product, accounting for sites that switched between products over time.

## Results

Across the 22 study districts, an average of 51 months (range 38-62) of organophosphate IRS (OP-IRS) data and 40 months (range 22-57) of neonicotinoid IRS (NN-IRS) data were included in the analysis (S2 Table). Overall, the observed malaria incidence averaged 233 per 1,000 person-years (range 75-410) during the OP-IRS period, compared to 298 per 1,000 person-years (range 95-447) during the NN-IRS period. Across most districts (18 of the 22 IRS districts), absolute malaria incidence was higher during the NN-IRS period compared to the OP-IRS period. The relative increases were most pronounced in Wa West, Lambussie, Lawra, and Daffiama-Bussie-Issa in the Upper West Region (Fig 2 & S2 Fig). Reductions in absolute malaria incidence were observed in only two districts - Wa Municipal (Upper West Region) and Kumbungu (Northern Region), while no change was recorded in Kassena-Nankana West and Mamprugu-Moagduri.

**Fig 2.**
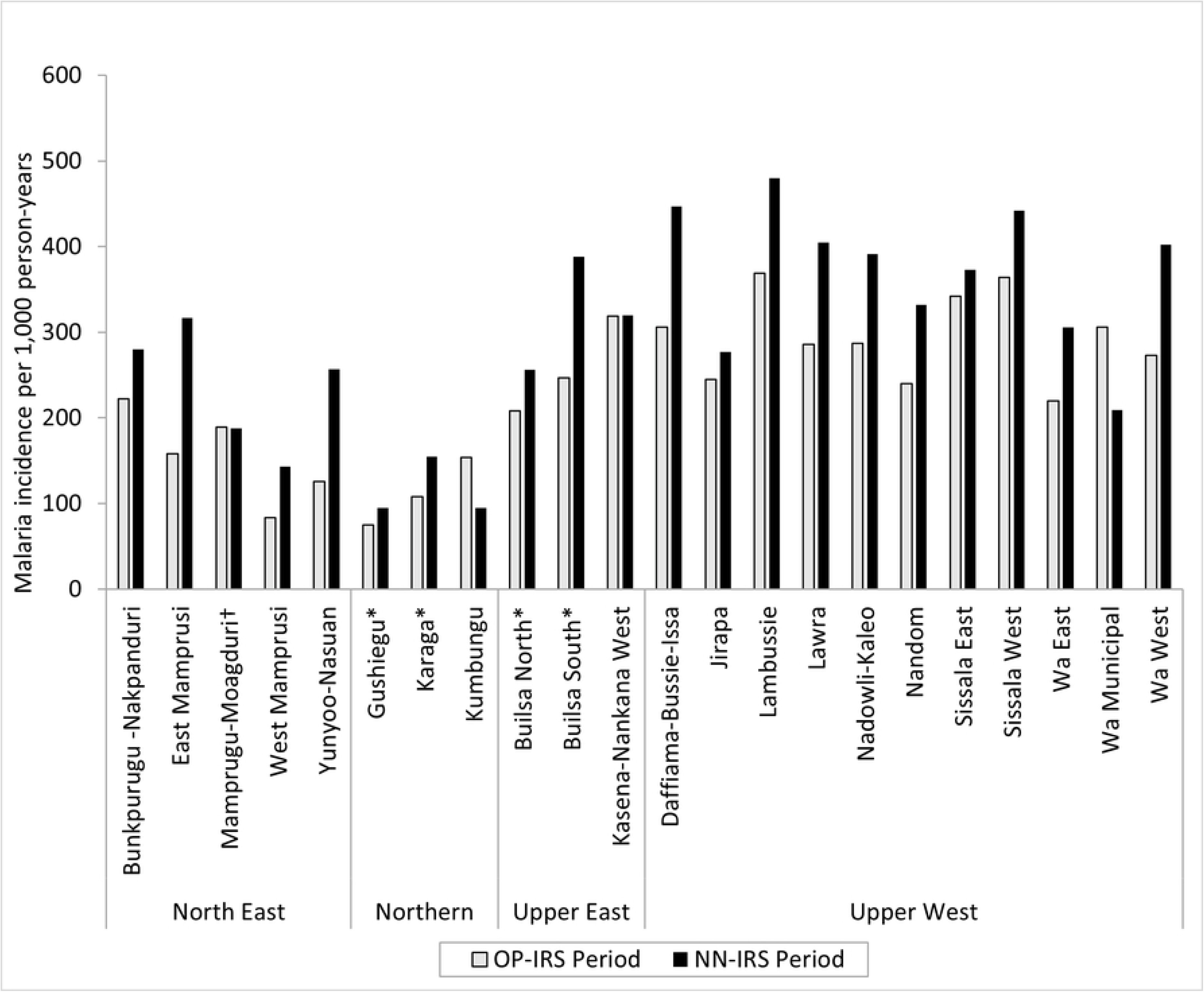
Comparison of mean absolute malaria incidence (per 1,000 person-years) across districts, grouped by region during periods when indoor residual spraying (IRS) was conducted with organophosphate (OP) insecticides versus neonicotinoids (NN). *Each pair of bars represents one district, with incidence rates from the OP-IRS period shown in grey and those from the NN-IRS period shown in black bars.* \**Observed start date for the period under consideration was January 2015, with an end date of December 2022 for all districts except Gushiegu, Karaga, Builsa North, Builsa South, and Kasena-Nankana West, for which the observed start date was January 2017*.

Observed malaria incidence across all regions, stratified by region, showed marked seasonal peaks (Fig 3). Results from the ITS analysis shows that in the months following the insecticide switch, observed incidence consistently exceeded the modelled counterfactual, which represents the expected incidence had organophosphate-based IRS been maintained. The divergence between observed and counterfactual trends was evident in the first transmission season post-switch and persisted across subsequent campaigns.

**Fig 3.**
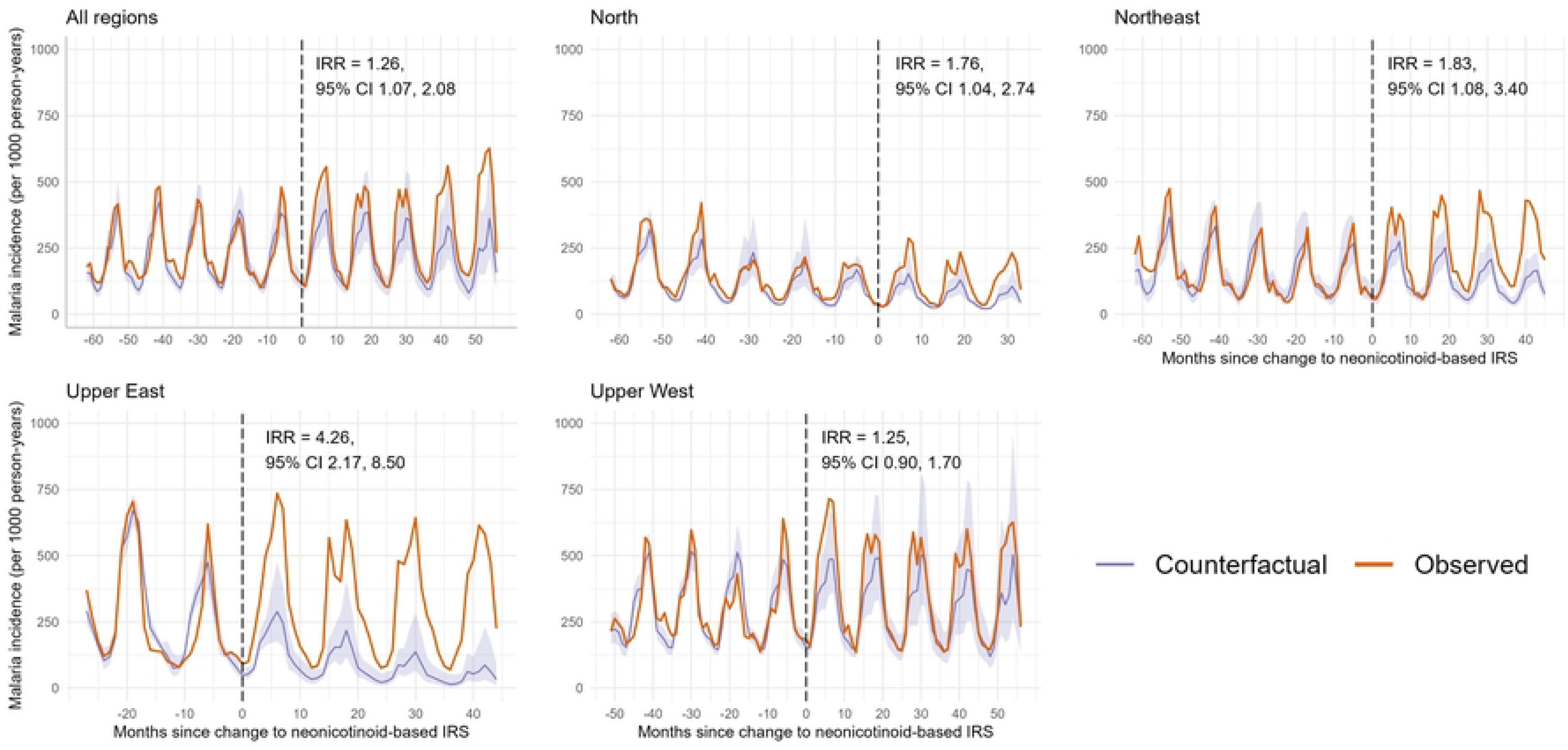
Observed and modelled counterfactual monthly malaria incidence before and after the change to neonicotinoid-based IRS, overall and stratified by region. *Panels show observed (orange) and modelled counterfactual (purple with 95% CI) monthly malaria incidence per 1,000 person-years pooled across All Regions, as well as the individual regions (Northern, North East, Upper East, and Upper West). The dashed line at Month 0 marks the switch from organophosphate to neonicotinoid IRS. “Observed” refers to the actual reported malaria incidence from DHIMS2 data, while “counterfactual” represents expected incidence had organophosphate (pirimiphos methyl) IRS continued. IRRs (with 95% CIs) indicate the impact of the switch, adjusted for rainfall, temperature, NDVI, and initiation of seasonal malaria chemoprevention*.

For all regions combined, the estimated IRR for the period after the switch was 1.26 (95% CI: 1.07-2.08). Regional differences were apparent (three-way joint interaction p = 0.004). In the Northern, North East, and Upper East regions, incidence was significantly higher after the switch compared with the counterfactual (Northern: IRR = 1.76, 95% CI 1.04-2.74; North East: IRR = 1.83, 95% CI 1.08-3.40; Upper East: IRR = 4.26, 95% CI 2.17-8.50). In contrast, in the Upper West region no significant difference was detected (IRR = 1.25, 95% CI 0.90-1.70). When Mamprugu-Moagduri, where *Actellic® 300CS* was reintroduced in 2020, was excluded from the analysis, malaria incidence increased substantially, from an overall regional incidence rate ratio (IRR) of 1.38 (95% CI: 1.12–1.92) to 2.57 (95% CI: 1.97–4.00) in the North East region (Supplemental S1 Fig).

To investigate regional differences further, we assessed whether baseline transmission intensity modified the impact of switching to neonicotinoid-based IRS. The effect of the change differed by baseline incidence category (Fig 4, 3-way joint interaction p = 0.002). In districts with low baseline incidence, malaria incidence increased by 54% relative to the counterfactual after the switch (IRR = 1.54, 95% CI 1.14-2.23). In moderate-incidence districts, the increase was 20% (IRR = 1.20, 95% CI 1.02-1.43).

**Fig 4.**
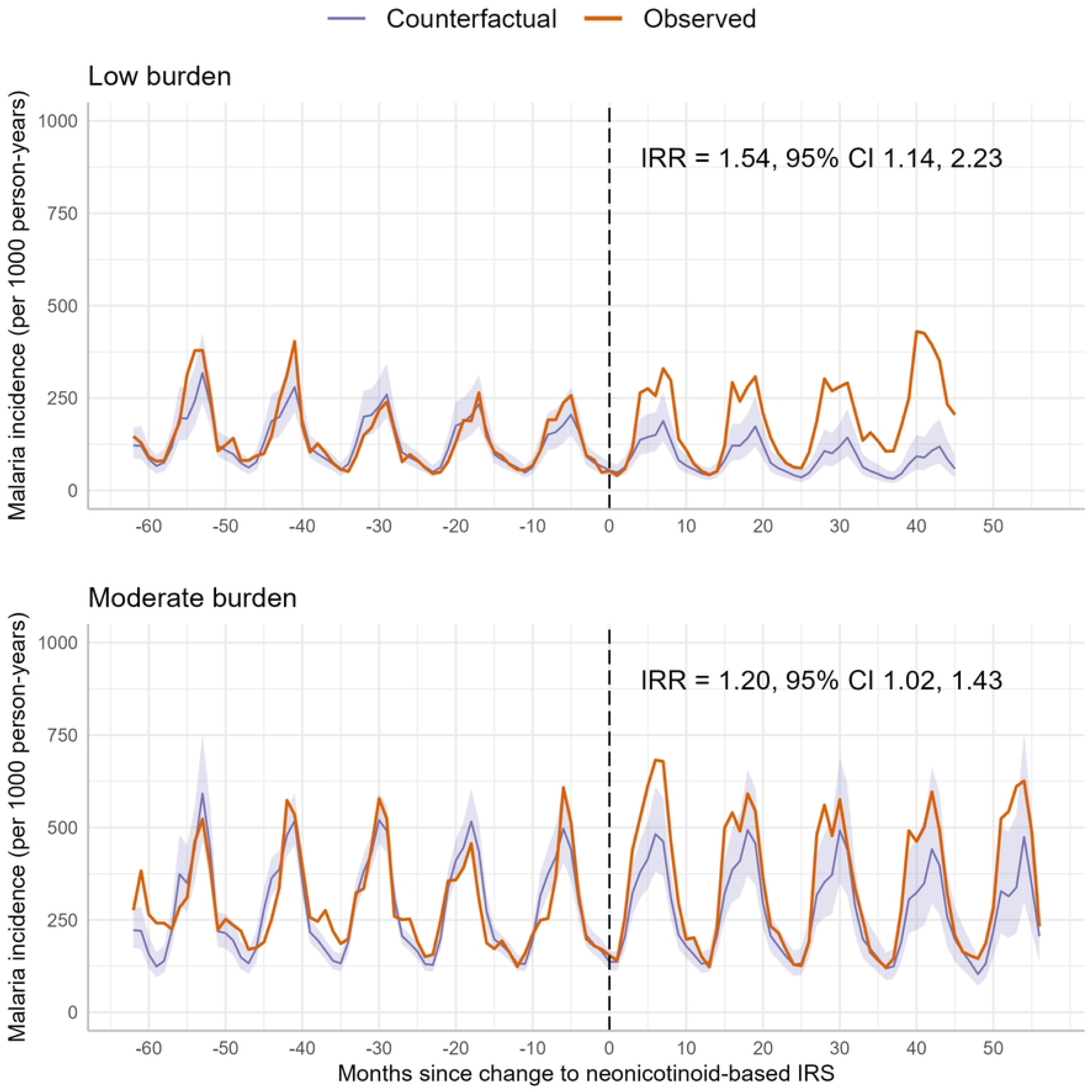
Observed and modelled counterfactual monthly malaria incidence before and after the change to neonicotinoid-based IRS, pooled and stratified by baseline malaria incidence. *This figure presents Monthly malaria incidence per 1,000 person-years before and after the switch to neonicotinoid IRS, stratified by baseline incidence (low, medium, high). Orange lines show observed DHIMS2 data; purple lines show counterfactual estimates with 95% CIs. Dashed line (Month 0) marks the switch. IRRs quantify post-switch changes relative to the counterfactual after adjusting for rainfall, temperature, and NDVI*.

In sensitivity analyses that examined whether the results differed by the two neonicotinoid-based IRS products (SumiShield vs. Fludora®Fusion), we found no evidence of a difference in immediate effect (χ²(1)=2.29, p=0.13) or in post-spray slope (χ²(1)=0.03, p=0.86) between the two formulations.

## Discussion

The interrupted time series revealed that replacing organophosphate insecticides with neonicotinoids in IRS campaigns across northern Ghana was associated with an overall 26% increase in malaria incidence, relative to a scenario in which organophosphates had been maintained. This was further confirmed with the re-introduction of organophosphate IRS in Mamprugu-Moagduri district resulting in a decline in cases. We found that the resurgence in malaria incidence was most pronounced in regions with low baseline malaria transmission, suggesting that neonicotinoid-based IRS might have been less effective in these settings compared to organophosphates. Particularly concerning was the Upper East region, where malaria incidence rose by 3.7 times the expected level in the three IRS-targeted districts following the transition to neonicotinoids. In contrast, areas with moderate malaria intensity showed a comparatively smaller increase, possibly due to already elevated transmission levels where marginal changes are less detectable. Although absolute malaria incidence rose in the Upper West region, where malaria intensity was highest, our analysis indicates that this trend cannot be attributed solely to the insecticide change. These findings underscore the critical need for robust, context-specific epidemiological monitoring when introducing new vector control products.

In our study we investigated the transition of insecticides in the same regions over time, with no indication that IRS acceptance, programmatic elements and vector-specific factors changed dramatically in this period [7, 46–49]. The spray coverages for the different IRS campaigns between 2015 and 2022 have ranged between 87% and 98% (above 80% target coverage [50, 51]) with high spray quality each year [19, 46, 52]. Vector species composition in the area has remained stable over the years with *An. gambiae* s.s. the predominant vector species, co-existing with *An. coluzzii* and *An. arabiensis* [19, 33, 53, 54]. Possible disruptions from the COVID-19 pandemic could have affected the provision and uptake of malaria prevention and treatment services, and might account for the increased malaria incidence in 2020 and 2021 compared to pre COVID-19 pandemic period, as has been reported in Uganda [22]. However, these disruptions are unlikely to account for the heterogeneous change in malaria incidence across the study area [39]. Notably, we recorded reductions in absolute malaria incidence in the Kumbungu district (Northern region) and Wa municipality (Upper West region), despite their similar IRS histories to other districts within their respective regions. Furthermore, the residual bioefficacy of both SumiShield 50WG and Fludora® Fusion have been reported to last between eight to nine months, which is similar to Actellic 300CS and exceed the duration of the malaria transmission season in the northern Savannah zone [19, 55]. The impact of neonicotinoids-based IRS on malaria incidence appears to be heterogeneous and is likely impacted by the simultaneous gradual implementation of SMC across the region. The overlap of the successful introduction and high uptake of SMC across the different regions with the change in IRS insecticides could indicate a possible bigger resurgence in malaria cases if SMC had not been introduced. Previous studies demonstrated the feasibility and effectiveness of SMC in reducing malaria incidence in target populations in Northern Ghana particularly when coverage is high[56]. SMC coverages in the study area ranged between 87% and 99% of the target populations [57–59].

The heterogeneity in the impact of the IRS insecticide change is likely also influenced by the susceptibility profile of the local *Anopheles* vector populations. Although the use of neonicotinoids for vector control was novel in Ghana at the time of introduction, this insecticide had already been widely used in agriculture in various formulations. In fact, as of November 2019, the Ghana Environmental Protection agency (EPA) had registered over 30 neonicotinoid-based products under different trade names for agricultural purposes [60]. Crops such as cotton, maize and soybean rely heavily on neonicotinoids-based pesticides to control common pests [60–62]. As these insecticides are highly water soluble, they readily leach into the soil and aquatic systems [63]. One study in 2015 found that 79% of the water samples in cotton growing communities in northern Ghana were positive for high concentration of neonicotinoids [64]. Given the life cycle of *Anopheles* mosquitoes in aquatic habitats, there is high likelihood of exposure to (sub-)lethal doses of neonicotinoids in areas where neonicotinoids are used in agriculture, as has been found in Cameroon [65, 66]. The limited impact of neonicotinoids on malaria incidence in parts of the northern Savannah zone coincides with areas of intensive cotton and soybean farming [67], suggesting potential cross-resistance due to agricultural use. Similar patterns are observed in IRS districts of Uganda, and Malawi involved in neonicotinoid-reliant agriculture [68, 69]. Further studies are needed to explore how agricultural insecticide exposure affects vector susceptibility and malaria control outcomes, including potential links between crop types and insecticide efficacy.

Baseline clothianidin susceptibility testing conducted in 2017 indicated that the malaria vectors in the study area were largely susceptible to clothianidin, even in high cotton production areas [49, 70], and this susceptibility appeared to persist after neonicotinoid IRS was introduced [37]. Yet, entomological surveillance after introduction of neonicotinoids showed high parity rates, indicating high survivorship of vector species. In the North-East IRS districts, parity rate rose from 37.6% during the organophosphate period to 45.2% under neonicotinoids (2020–2022), a 20% relative increase, suggesting a higher proportion of older, potentially infectious mosquitoes. Conversely, parity rates in unsprayed areas have shown consistent decline (from 69.0% to 58.2%) over the same period [37, 52]. These findings, alongside the increase in malaria incidence after neonicotinoid IRS, calls into question the operational sensitivity of current WHO phenotypic susceptibility tests [71], which may only detect intense resistance. Furthermore, the use of rapeseed oil-based surfactants in bottle bioassays may overestimate clothianidin potency [72, 73], possibly explaining the lack of detected resistance at baseline. Enhanced frequency and geographic targeting of resistance monitoring are needed, especially considering the overlap between agricultural pesticide use and IRS sites. This is particularly relevant as most of the newly approved IRS products, including Vectron T500 (a.i. broflanilide), Sylando 240SC (a.i. chlorfenapyr), and Sovrenta 15WP (a.i. Isocycloseram) [74], are repurposed agricultural insecticides [75–77].

While our study provides valuable insights, it is not without limitations. This study is based on passive surveillance data at district level, which may have introduced reporting biases, that could have influenced the modelled estimates. Health-seeking behaviour data were limited to a single DHS survey, restricting our ability to identify changes in behaviour and whether trends observed in passive case detection accurately reflect true malaria incidence. While there has been a general improvement in the DHIS2 data quality since 2014, there is the possibility of data quality issues-incomplete, missing data, over reporting, low or high testing, false positives could also bias the estimates. As data are aggregated to the district-level, important geographical variations might have been missed. The analysis itself used general factors to model malaria incidence. More studies on factors contributing to malaria incidence in the area are necessary to fully understand the relationship between variation in IRS activities and malaria incidence especially in the Upper West region. Additionally, qualitative studies exploring community coverages, perceptions, and adherence to IRS programs could provide a more comprehensive understanding of the factors influencing the effectiveness of malaria control interventions.

## Conclusion

This study highlights the importance of understanding the impact of switching to a new insecticides in IRS programs on malaria case incidence outcomes to inform decisions on malaria programming. Due to increasing insecticide resistance, many malaria programs have added neonicotinoid-based insecticides in their IRS programs for effective resistance management. Recent reports of resurgence in malaria incidence in Uganda and now Ghana following the transition of IRS insecticides to neonicotinoids raises questions about long-term sustainability of neonicotinoids for IRS in areas where neonicotinoids are used extensively for agriculture. Our findings highlight the need for more studies on the reasons why neonicotinoids are underperforming compared to organophosphates in certain environments and underscores the importance of tailoring malaria control strategies to specific regional contexts using detailed resistance profiles and deeper understanding of the impact of agricultural pesticides. Malaria control programs will need to consider a more tailored strategy, integrating multiple control methods and varying insecticides based on subnational vector resistance profiles, ecological conditions and epidemiological outcomes.

## Data Availability

All data supporting the conclusions of this article are included within the article and supplementary files.

## Acknowledgments

We would like to thank the U.S President’s Malaria Initiative and Abt Associates (through the VectorLink Ghana Project, now PMI Evolve), the Global Fund to fight AIDS, Tuberculosis and Malaria, and AngloGold Ashanti for the funding support for IRS in Ghana. We are also very grateful to the National Malaria Elimination Programme of Ghana, the regional and district staff of the Ghana Health Services for their support of this work. The critical feedback and inputs from the PMI Evolve Ghana team and AGAMal Ltd. is also greatly appreciated.

## Author Contributions

**Conceptualization:** Sylvester Coleman, Keziah Malm, Christian Atta-Obeng, Otubea Owusu Akrofi, Julie-Anne A Tangena

**Data curation:** Sylvester Coleman, Abdul Gafaru Mohammed, Ihsan Isaka

**Formal analysis:** Sylvester Coleman, Adrienne Epstein, Julie-Anne A Tangena, Abdul Gafaru Mohammed

**Data Validation:** Abdul Gafaru Mohammed, Ihsan Isaka, Wahjib Mohammed, Nana Yaw Peprah

**Visualization:** Adrienne Epstein, Clinton Nkolokosa.

**Writing – original draft:** Sylvester Coleman, Julie-Anne A Tangena

**Writing – review & editing:** Sylvester Coleman, Christian Atta-Obeng, Clinton Nkolokosa, Abdul Gafaru Mohammed, Otubea Owusu Akrofi, Ihsan Isaka, Wahjib Mohammed, Nana Yaw Peprah, Samuel Asiedu, Samuel K Dadzie, Dominic Dery, Julie-Anne A Tangena, Adrienne Epstein, Keziah Malm.

## Supporting information

**S1 Fig. Results from interrupted time series analysis when leaving out one district (Mamprugu-Moagduri) that reintroduced organophosphates in 2020.**

**S2 Fig. District-level percentage change in malaria incidence following neonicotinoid IRS in northern Ghana.**

**S1 Table. Summary of type of insecticides used in our study districts from 2008-2022.**

**S2 Table. Pre-and Post-Neonicotinoid IRS Malaria Incidence grouped by districts and region**

**S3 Table. Incidence rate ratios comparing observed malaria incidence to counterfactual malaria incidence modelled using interrupted time series methods**

## References

1. WHO. Global technical strategy for malaria 2016-2030: World Health Organization; 2015.

2. WHO Fa. International Code of Conduct on Pesticide Management – Guidance on good labelling practice for pesticides. Second revision ed. Rome, Italy 2022.

3. Bhatt S, Weiss D, Cameron E, Bisanzio D, Mappin B, Dalrymple U, et al. The effect of malaria control on Plasmodium falciparum in Africa between 2000 and 2015. Nature. 2015;526(7572):207–11.

4. Cibulskis RE, Alonso P, Aponte J, Aregawi M, Barrette A, Bergeron L, et al. Malaria: global progress 2000–2015 and future challenges. Infectious diseases of poverty. 2016;5(1):1–8.

5. Yukich J, Digre P, Scates S, Boydens L, Obi E, Moran N, et al. Incremental cost and cost-effectiveness of the addition of indoor residual spraying with pirimiphos-methyl in sub-Saharan Africa versus standard malaria control: results of data collection and analysis in the Next Generation Indoor Residual Sprays (NgenIRS) project, an economic-evaluation. Malaria journal. 2022;21(1):185. doi: 10.1186/s12936-022-04160-3.

6. Wagman J, Gogue C, Tynuv K, Mihigo J, Bankineza E, Bah M, et al. An observational analysis of the impact of indoor residual spraying with non-pyrethroid insecticides on the incidence of malaria in Ségou Region, Mali: 2012-2015. Malaria journal. 2018;17(1):19. doi: 10.1186/s12936-017-2168-2.

7. Gogue C, Wagman J, Tynuv K, Saibu A, Yihdego Y, Malm K, et al. An observational analysis of the impact of indoor residual spraying in Northern, Upper East, and Upper West Regions of Ghana: 2014 through 2017. Malaria journal. 2020;19(1):242. doi: 10.1186/s12936-020-03318-1.

8. Akogbéto MC, Aïkpon RY, Azondékon R, Padonou GG, Ossè RA, Agossa FR, et al. Six years of experience in entomological surveillance of indoor residual spraying against malaria transmission in Benin: lessons learned, challenges and outlooks. Malaria Journal. 2015;14(1):242.

9. Aïkpon R, Sèzonlin M, Tokponon F, Okè M, Oussou O, Oké-Agbo F, et al. Good performances but short lasting efficacy of Actellic 50 EC Indoor Residual Spraying (IRS) on malaria transmission in Benin, West Africa. Parasites & vectors. 2014;7(1):1–8.

10. WHO. World malaria report 2022. Geneva: World Health Organization; 2022.

11. WHO. World malaria report 2023. Geneva: World Health Organization; 2023.

12. Kleinschmidt I, Bradley J, Knox TB, Mnzava AP, Kafy HT, Mbogo C, et al. Implications of insecticide resistance for malaria vector control with long-lasting insecticidal nets: a WHO-coordinated, prospective, international, observational cohort study. The Lancet infectious diseases. 2018;18(6):640–9.

13. Kleinschmidt I, Bradley J, Knox TB, Mnzava AP, Kafy HT, Mbogo C, et al. Implications of insecticide resistance for malaria vector control with long-lasting insecticidal nets: a WHO-coordinated, prospective, international, observational cohort study. The Lancet Infectious diseases. 2018;18(6):640–9. doi: 10.1016/S1473-3099(18)30172-5.

14. Tangena J-AA, Hendriks CMJ, Devine M, Tammaro M, Trett AE, Williams I, et al. Indoor residual spraying for malaria control in sub-Saharan Africa 1997 to 2017: an adjusted retrospective analysis. Malaria journal. 2020;19(1):150. doi: 10.1186/s12936-020-03216-6.

15. WHO. Report of the sixteenth WHOPES working group meeting: WHO/HQ, Geneva, 22-30 July 2013: review of Pirimiphos-methyl 300 CS, Chlorfenapyr 240 SC, Deltamethrin 62.5 SC-PE, Duranet LN, Netprotect LN, Yahe LN, Spinosad 83.3 Monolayer DT, Spinosad 25 Extended release GR. Geneva: WHO, 2013 924150630X Contract No.: WHO/HTM/NTD/WHOPES/2013.6.

16. Oxborough RM. Trends in US President’s Malaria Initiative-funded indoor residual spray coverage and insecticide choice in sub-Saharan Africa (2008–2015): urgent need for affordable, long-lasting insecticides. Malaria journal. 2016;15(1):146.

17. WHO. Prequalification Vector Control: SumiShield 50WG 2017 [cited 2020 2 November 2020]. Available from: http://158.232.12.119/pq-vector-control/prequalified-lists/sumishield50wg/en/.

18. WHO. Prequalification Vector Control: Fludora Fusion 2018 [cited 2020 2 November 2020]. Available from: https://www.who.int/pq-vector-control/prequalified-lists/FludoraFusion/en/.

19. PMI/VL. The PMI VectorLink Project. March 2022. Annual Entomological Monitoring Report for Northern Ghana, March 1–December 31, 2021. Rockville, MD: USA: Abt Associates Inc., 2022 March 2022. Report No.

20. Odjo EM, Akpodji CST, Djènontin A, Salako AS, Padonou GG, Adoha CJ, et al. Did the prolonged residual efficacy of clothianidin products lead to a greater reduction in vector populations and subsequent malaria transmission compared to the shorter residual efficacy of pirimiphos-methyl? Malaria journal. 2024;23(1):119. doi: 10.1186/s12936-024-04949-4.

21. Emily RH, Ndombour G-C, Auguste A, Mathieu E, John K, Barthelemy G, et al. Reduction of malaria case incidence following the introduction of clothianidin-based indoor residual spraying in previously unsprayed districts: an observational analysis using health facility register data from Côte d’Ivoire, 2018-2022. BMJ Global Health. 2024;9(3):e013324. doi: 10.1136/bmjgh-2023-013324.

22. Epstein A, Maiteki-Sebuguzi C, Namuganga JF, Nankabirwa JI, Gonahasa S, Opigo J, et al. Resurgence of malaria in Uganda despite sustained indoor residual spraying and repeated long lasting insecticidal net distributions. PLOS Global Public Health. 2022;2(9):e0000676.

23. NMCP. Integrated Malaria Vector Management Policy. 2020 Edition ed. Accra Ghana: Ministry of Health, Ghana Health Services 2020. 60 p.

24. PMI. President’s Malaria Initaitive Ghana Malaria Operational Plan – FY08 President’s Malaria Initaitive 2007.

25. PMI. U.S. President’s Malaria Initiative Ghana Malaria Operational Plan FY 2020. 2019.

26. PMI|VectorLink. Ghana End of Spray Report: Spray Campaign: March 24-April 28, 2020. Rockville, MD, USA: The PMI VectorLink Project, Abt Associates Inc., 2020 June 2020. Report No.

27. Gogue C, Wagman J, Tynuv K, Saibu A, Yihdego Y, Malm K, et al. An observational analysis of the impact of indoor residual spraying in Northern, Upper East, and Upper West Regions of Ghana: 2014 through 2017. Malaria journal. 2020;19(1):1–13.

28. Sherrard-Smith E, Jamie GT, Winskill P, Corbel V, Pennetier C, Djénontin A, et al. Systematic review of indoor residual spray efficacy and effectiveness against *Plasmodium falciparum* in Africa. Nature Communications. 2018;9 (1)(4982).

29. Owusu K. Rainfall changes in the savannah zone of northern Ghana 1961–2010. Weather. 2018;73(2):46–50.

30. Manzoor SA, Griffiths GH, Robinson E, Shoyama K, Lukac M. Linking pattern to process: intensity analysis of land-change dynamics in Ghana as correlated to past socioeconomic and policy contexts. Land. 2022;11(7):1070.

31. Survey USG. Landsat Surface Reflectance-derived Spectral Indices 2023. Available from: https://www.usgs.gov/landsat-missions/landsat-surface-reflectance-derived-spectral-indices.

32. PMI. Ghana Malaria Profile. US President’s Malaria Initiative, 2023.

33. PMI/VL. The PMI VectorLink Project. Annual Entomological Monitoring Report for Northern Ghana, March 1-December 31, 2018. Abt Associates Inc, 2019 March 2019. Report No.

34. Gakpey K, Baffoe-Wilmot A, Malm K, Dadzie S, Bart-Plange C. Strategies towards attainment of universal coverage of long lasting insecticide treated nets (LLINs) distribution: experiences and lessons from Ghana. Parasites & vectors. 2016;9(1):35.

35. Kumah E, Duvor F, Otchere G, Ankomah SE, Fusheini A, Kokuro C, et al. Intermittent preventive treatment of Malaria in pregnancy with sulphadoxine-pyrimethamine and its Associated factors in the Atwima Kwanwoma District, Ghana. Annals of global health. 2022;88(1):27.

36. PMI. U.S. President’s Malaria Initiative Ghana Malaria Operational Plan FY 2023 Retrieved from www.pmi.gov. 2023.

37. PMI/VL. The PMI VectorLink Project Annual Entomological Monitoring Report for Northern Ghana, March 1–December 31, 2022. Rockville, Maryland: Abt Associates Inc. 2023.

38. GSS. Ghana Multiple Indicator Cluster Survey with an Enhanced Malaria Module and Biomarker, 2011, Final Report. Ghana Statistical Service Accra, Ghana., 2011.

39. ICF Ga. Ghana Demographic and Health Survey 2022: Key Indicators Report. Accra, Ghana: Ghana Statistical Services, Accra Ghana and The DHS Program ICF Rockville, Maryland, USA, 2023.

40. Ampofo GD, Osarfo J, Aberese-Ako M, Asem L, Komey MN, Mohammed W, et al. Malaria in pregnancy control and pregnancy outcomes: a decade’s overview using Ghana’s DHIMS II data. Malar J. 2022;21(1):303. Epub 2022/10/29. doi: 10.1186/s12936-022-04331-2. PubMed PMID: 36303165; PubMed Central PMCID: PMCPMC9615308.

41. Abatzoglou J. Terraclimate, a high-resolution global dataset of monthly climate and climatic water balance. Available from: https://www.climatologylab.org/terraclimate.html.

42. Center CH. CHIRPS-2.0 2023. Available from: https://data.chc.ucsb.edu/products/CHIRPS-2.0/.

43. Bernal JL, Cummins S, Gasparrini A. Interrupted time series regression for the evaluation of public health interventions: a tutorial. International journal of epidemiology. 2017;46(1):348–55.

44. Kontopantelis E, Doran T, Springate DA, Buchan I, Reeves D. Regression based quasi-experimental approach when randomisation is not an option: interrupted time series analysis. bmj. 2015;350.

45. Højsgaard S, Halekoh U, Yan J. The R package geepack for generalized estimating equations. Journal of statistical software. 2006;15:1–11.

46. PMI|AIRS. Entomological monitoring of the PMI AIRS program in Northern Ghana. 2017 Annual Report. President’s Malaria Initiative/Africa Indoor Residual Spraying Program. Abt Associates Inc., 2017

47. PMI/AIRS. Entomological Monitoring of the PMI AIRS program in Northern Ghana. 2016 Annual Report, Bethesda, MD, Abt Associates Inc. PMI | Africa IRS (AIRS) Project Indoor Residual Spraying, 2016.

48. PMI/VL. The PMI VectorLink Project Annual Report: October 1, 2020–September 30, 2021. Rockville, Maryland: The PMI VectorLink Project, Abt Associates. 2021 November, 2021. Report No.

49. PMI/VL. The PMI VectorLink Project. November 2022. The PMI VectorLink Project Annual Report: October 1, 2021–September 30, 2022. The PMI VectorLink Project, Abt Associates,Rockville, Maryland, 2022 2022. Report No.

50. Rehman AM, Coleman M, Schwabe C, Baltazar G, Matias A, Gomes IR, et al. How much does malaria vector control quality matter: the epidemiological impact of holed nets and inadequate indoor residual spraying. PloS one. 2011;6(4):e19205.

51. WHO. Operational manual on indoor residual spraying: control of vectors of malaria, Aedes-borne diseases, Chagas disease, leishmaniases and lymphatic filariasis. Geneva2023. Available from: https://www.who.int/publications/i/item/9789240083998.

52. PMI/VL. The PMI VectorLink Project Annual Report: October 1, 2018-September 30, 2019. Rockville, Maryland.: The PMI VectorLink Project,Abt Associates Inc, 2019.

53. Coleman S, Dadzie S, Aklilu S, Mumba P, Yihdego Y, Dengela D, et al. A reduction in malaria transmission intensity in Northern Ghana after 7 years of indoor residual spraying. Malaria journal. 2017:1–10.

54. Appawu M, Owusu-Agyei S, Dadzie S, Asoala V, Anto F, Koram K, et al. Malaria transmission dynamics at a site in northern Ghana proposed for testing malaria vaccines. Tropical Medicine & International Health. 2004;9(1):164–70.

55. PMI/VL. The PMI VectorLink Project. Annual Entomological Monitoring Report for Northern Ghana, March 1-December 31, 2018. Abt Associates Inc, 2019 March 2019. Report No.

56. Ansah PO, Ansah NA, Malm K, Awuni D, Peprah N, Dassah S, et al. Evaluation of pilot implementation of seasonal malaria chemoprevention on morbidity in young children in Northern Sahelian Ghana. Malaria journal. 2021;20(1):440. doi: 10.1186/s12936-021-03974-x.

57. DHIS2 G. Ghana Health Service District Health Information System 2 (DHIS-2) database, 2012– 2022. 2022.

58. Ayamba EY, Dzotsi EK, Dormechele W, Ansah NA, Bangre O, Nyuzaghl JA-I, et al. Evaluation of seasonal malaria chemoprevention implementation in the Upper East region of Northern Ghana. Malaria journal. 2025;24(1):123. doi: 10.1186/s12936-025-05322-9.

59. NMCP. Synopsis for 2022 World Malaria Day Commemoration. Accra Ghana: National Malaria Control Proramme Post Office Box KB 493, Korle Bu, Accra, 2022.

60. USAID|Ghana. Review of Pesticide Evaluation Report and Safe Use and Action Plan (PERSUAP) for USAID Funded Feed The Future Ghana Agricultural Development And Value Chain Enhancement Project (FTF ADVANCE II) 2019.

61. Williams F, Marwijk Av, Rware H, Essegbey G, Besah P, Duah S, et al. Implementation of Fall Armyworm Management Plan in Ghana. Plant Health Cases. 2023;(2023):phcs20230013.

62. Bariw SA, Kudadze S, Adzawla W. Prevalence, effects and management of fall army worm in the Nkoranza South Municipality, Bono East region of Ghana. Cogent Food & Agriculture. 2020;6(1):1800239.

63. Africa AoSoS. Neonicotinoid Insecticides: Use and Effects in African Agriculture: A Review and Recommendations to Policymakers. 2021.

64. Abdulahi A, Obiri-Danso K, Mohammed S. Effect of farmers’ attitude, usage pattern and handling of pesticides on potable water quality in northern Ghana. International Journal of Development and Sustainability. 2015;Volume 4 Number 10 (2015): 977–87.

65. Tchouakui M, Assatse T, Mugenzi LMJ, Menze BD, Nguiffo-Nguete D, Tchapga W, et al. Comparative study of the effect of solvents on the efficacy of neonicotinoid insecticides against malaria vector populations across Africa. Infectious diseases of poverty. 2022;11(1):35. doi: 10.1186/s40249-022-00962-4.

66. Nkya TE, Akhouayri I, Poupardin R, Batengana B, Mosha F, Magesa S, et al. Insecticide resistance mechanisms associated with different environments in the malaria vector *Anopheles gambiae*: a case study in Tanzania. Malaria journal. 2014;13(1):1–15.

67. USDA. West Africa - Crop Production Maps: U.S. Department of Agriculture 2023. Available from: https://ipad.fas.usda.gov/rssiws/al/wafrica_cropprod.aspx.

68. USDA. Southern Africa - Crop Production Maps: U.S. Department of Agriculture 2023. Available from: https://ipad.fas.usda.gov/rssiws/al/safrica_cropprod.aspx.

69. USDA. East Africa - Crop Production Maps: U.S. Department of Agriculture 2023. Available from: https://ipad.fas.usda.gov/rssiws/al/eafrica_cropprod.aspx.

70. PMI/VL. The PMI VectorLink Project. PMI VectorLink Malawi Annual Entomological Monitoring Report, July 1, 2021 – June 30, 2022. Rockville, MD: USA: Abt Associates Inc., 2022 September 2022. Report No.

71. Corbel V, Kont MD, Ahumada ML, Andréo L, Bayili B, Bayili K, et al. A new WHO bottle bioassay method to assess the susceptibility of mosquito vectors to public health insecticides: results from a WHO-coordinated multi-centre study. Parasites & vectors. 2023;16(1):21. doi: 10.1186/s13071-022-05554-7.

72. Ashu FA, Fouet C, Ambadiang MM, Penlap-Beng V, Kamdem C. Vegetable oil surfactants are synergists that can bias neonicotinoid susceptibility testing in adult mosquitoes. 2023. doi: 10.1101/2023.04.18.537421.

73. WHO. Quality of conduct and interpretation of insecticide susceptibility in bioassays using new insecticides for vector control: meeting report,. Geneva: World Health Organization, 2025 24 September 2024. Report No.

74. WHO. Vector Control Products Pipeline: World Health Organization; 2023. Available from: https://extranet.who.int/prequal/vector-control-products/prequalification-pipeline.

75. van Herk WG, Vernon RS, Goudis L, Mitchell T. Broflanilide, a meta-diamide insecticide seed treatment for protection of wheat and mortality of wireworms (Agriotes obscurus) in the field. Journal of Economic Entomology. 2021;114(1):161–73.

76. EPA. Pesticide Fact Sheet Chlorfenapyr Reason for Issuance: New Chemical Registration. 2001 January, 2001. Report No.: Contract No.: EPA-730-F-00-001.

77. van Herk WG, Vernon RS, Labun T, Spies J. Isocycloseram, a novel isoxazoline insecticide seed treatment for protection of wheat and barley and mortality of wireworms, Limonius californicus (Coleoptera: Elateridae). J Econ Entomol. 2024;117(5):1926–37. Epub 2024/07/31. doi: 10.1093/jee/toae170. PubMed PMID: 39082981.

